# Prevalence and factors associated with peripheral artery disease among patients with diabetes mellitus: A cross-sectional study at a tertiary hospital in Eastern Uganda

**DOI:** 10.64898/2026.06.03.26354843

**Authors:** Julius Imalingat, Asad Muyinda, Daniel Iraguha, Richard Katuramu, Peter M. Masaba, Esther Apio, Jasper Kebesu, Oreb Nankunda, Erias Kirabo, Joshua Epuitai, Denis Bwayo

## Abstract

**Background:** Peripheral artery disease (PAD) is a major contributor to morbidity and mortality, particularly among individuals with diabetes mellitus (DM), in whom its prevalence is markedly increased. PAD is often asymptomatic and under-diagnosed, especially in low-resource settings. This study aimed to determine the prevalence of PAD and associated factors among adults with DM in Eastern Uganda.

**Methods:** We conducted a hospital-based cross-sectional study at Mbale Regional Referral Hospital from 10^th^/12/2024 to 30^th^/04/2025. A total of 300 adult patients with DM were consecutively enrolled. Data on sociodemographic characteristics, clinical characteristics, comorbidities, and behavioural risk factors were collected using an interviewer-administered data tool. PAD was assessed using the ankle–brachial index (ABI), defined as ≤ 0.90. Modified Poisson regression was used to identify factors associated with PAD. As a secondary measure for PAD, we administered the Edinburgh Claudication Questionnaire (ECQ) to capture symptomatic PAD.

**Results:** The majority of the participants had a low fruit intake (68%), physical inactivity (54%), and elevated low-density lipoprotein (60%). The prevalence of PAD as measured by ABI was 42.3% (127/300; 95% CI 0.38-0.48), while the magnitude of PAD as measured by ECQ, combining participants with possible claudication and definite claudication was 37.3% 95% CI 31.9 - 42.8). Out of participants with PAD, 15.8% (20/127) were classified as having severe PAD (ABI <0.4). Socio-demographic and clinical factors were assessed for association with PAD. We found no evidence of association between the examined factors and PAD. Age (aPR 1.24 95% CI: 0.73 - 2.09), female sex (aPR 1.46 95% CI: 0.84 - 2.55), cholesterol level (aPR 1.39 95% CI: 0.86 - 2.25), glycemic control (aPR 1.35 95% CI: 0.72 - 2.53), and sedentary behaviour (aPR 1.28 95% CI: 0.79-2.08).

**Conclusion:** The prevalence of PAD was high among adults with DM in Eastern Uganda. Routine health education, and ABI screening of PAD should be done for patients living with DM. The absence of significant associations despite high prevalence of PAD may reflect unmeasured factors e.g. chronic inflammation that may be unique to this population, future prospective studies with larger sample size and more detailed objective measures e.g. inflammatory markers are needed to determine locally relevant modifiable risk factors.

## Background

Peripheral artery disease (PAD) ) is partial or complete atherosclerotic obstruction of the arteries supplying the lower extremities [1]. As a manifestation of systemic atherosclerotic disease, PAD is an established predictor of major cardiovascular events, including stroke and myocardial infarction [2, 3]. Among individuals with diabetes mellitus (DM), PAD contributes significantly to complications such as foot ulceration, which affects an estimated 9–26 million people annually. PAD is associated with reduced quality of life, increased healthcare utilization, and loss of productivity [4, 5].

Globally, approximately 202 million people are living with PAD. The burden of PAD continues to rise, largely driven by the increasing prevalence of non-communicable diseases such as DM [6-8]. Individuals with DM are at a two-to four-fold higher risk of developing PAD compared to those without diabetes [2, 9, 10]. In sub-Saharan Africa (SSA), available evidence suggests a high burden of PAD, particularly among older adults and individuals with DM [11]. In northwest Ethiopia, a prevalence of 30% was reported among patients with type 2 DM [9, 10, 12]. In Uganda, reported prevalence estimates range from 24% to 39%, reflecting variability across study populations and settings [10, 13].

Several modifiable and non-modifiable factors have been associated with PAD. Increasing age, particularly above 60 years, has consistently been identified as a major factor associated with PAD [12, 14, 15]. While some studies report a male predominance [9, 10, 14], others highlight the role of clinical and metabolic factors, including poor glycemic control, longer duration of diabetes, hypertension, dyslipidemia, and obesity [8, 12, 15]. Behavioral factors such as smoking, alcohol use, and physical inactivity have also been implicated, with smoking associated with a two-to five-fold increased risk of PAD [2, 8-10, 12, 15].

Despite its clinical importance, PAD remains underdiagnosed, largely because many patients are asymptomatic or present with atypical symptoms. In Uganda, evidence on PAD among individuals with DM is limited. To date, a paucity of studies have been conducted and the few were more than a decade ago in Western and Central Uganda that examined its prevalence and associated factors [9, 10]. The variability in reported prevalence and differences in study settings underscore the need for updated, context-specific data. This study was therefore conducted to determine the prevalence of PAD and associated factors among adults with DM attending a tertiary hospital in Eastern Uganda.

## Methods

### Study design, site, and population

This was a cross-sectional study conducted at the diabetic outpatient clinic of Mbale Regional Referral Hospital between 10^th^/12/2024 and 30^th^/04/2025. The hospital is located approximately 222 km east of Kampala in Mbale City, Eastern Uganda. It is one of 16 regional referral hospitals within the Ugandan public health system. The hospital functions as a tertiary care center serving a catchment population of approximately 4.5 million people. The diabetic clinic operates once a week with an average attendance of 100–150 patients per day including approximately 5–10 patients newly diagnosed with DM. Care is delivered by a multidisciplinary team comprising of physicians, internal medicine residents, medical officers, and nurses. The study population included adult patients aged 18 years and above with a confirmed diagnosis of diabetes mellitus who attended the outpatient diabetic clinic during the study period. Patients with bilateral lower limb amputation and those who were critically ill were excluded from the study.

### Sample size and sampling procedure

The sample size was calculated using the Kish–Leslie formula for cross-sectional studies. The prevalence of PAD of 24% from a similar population in western Uganda was used for determining the sample size [16]. This yielded a minimum required sample size of 291 participants. To account for potential nonresponse or incomplete data, the final sample size was increased to 300 participants. Participants were enrolled consecutively based on their order of attendance at the diabetic clinic until the required sample size was attained, with a daily recruitment target of approximately 25 participants. To prevent duplicate enrollment, each participant was assigned a unique identification code recorded on their appointment card. This identifier was verified at subsequent visits to ensure that no individual was enrolled more than once during the study period. The sampling process was conducted in close coordination with clinic staff to ensure seamless integration with routine clinical activities, without disrupting patient care.

### Study Variables

The primary outcome of interest was the presence of peripheral arterial disease (PAD), defined as an abnormal ankle–brachial index (ABI) ≤ 0.90. ABI was determined by measuring systolic blood pressure (SBP) at the brachial and ankle levels using 8 MHz handheld vascular Doppler device (Elite model No. 100) and an appropriately sized blood pressure cuff. The patient was asked to lie comfortably on an examination bed in the supine position. Any tightly fitting clothing was removed, and the patient was allowed to rest and remain relaxed for ten minutes before blood pressure measurements were taken by a trained research assistant who was a medical doctor. Brachial SBP was measured in both arms, and the higher of the two readings was used for ABI calculation. Ankle SBP was measured in both lower limbs at the posterior tibial and dorsalis pedis arteries. For each limb, the higher of the two ankle pressures was recorded. The ABI for each limb was calculated as the ratio of ankle SBP (either posterior tibial or dorsalis pedis, whichever was higher) to the higher brachial SBP. The participant’s overall ABI was defined as the lower value of the left and right limb ABIs. ABI values were categorized as follows: normal (≥ 0.91), mild PAD (0.70–0.90), moderate PAD (0.40–0.69), and severe PAD (< 0.40) [17].

As a screening approach, PAD was assessed using the ECQ (Appendix 1). Both the ABI and the ECQ were used as complementary screening tools to enhance the internal validity of PAD detection by integrating objective hemodynamic assessment with a standardized symptom-based evaluation. The ECQ is a validated screening tool that assesses symptoms of intermittent claudication through a structured history focused on exertional lower limb pain and its relief with rest. Based on participants’ responses, individuals were classified into three categories: definite claudication, possible claudication, or no claudication.

Independent variables encompassed a range of sociodemographic, clinical, lifestyle, anthropometric, and laboratory factors. Sociodemographic characteristics included age, sex, ethnicity, marital status, level of education, and occupation. Clinical variables comprised duration of diabetes mellitus (DM), and comorbid conditions such as hypertension and chronic kidney disease. Lifestyle-related factors included history of cigarette smoking, physical inactivity, and low consumption of fruits and vegetables. Anthropometric measurements included height, weight, waist circumference, and waist-to-height ratio. A 12 lead Electrocardiogram (ECG) was done for all patients. Laboratory parameters assessed were triglycerides, total cholesterol, high-density lipoprotein (HDL), low-density lipoprotein (LDL), fasting blood glucose (FBG), and glycated hemoglobin (HbA1c).

### Data Collection Tool and procedure

Data were collected using a pretested, structured questionnaire administered by trained research assistants under the supervision of the principal investigator. Prior to study initiation, all research assistants underwent comprehensive training on study procedures to ensure standardization and adherence to the study protocol. Written informed consent was obtained from all participants or their caregivers before commencement of any study-related procedures.

Sociodemographic information was obtained using a standardized data collection tool and included age, sex, ethnicity, religion, marital status, highest level of education attained, and occupation. A focused medical history was taken to ascertain risk factors for PAD, including history of cigarette smoking, physical inactivity, duration of diabetes mellitus, presence of comorbidities (such as hypertension and kidney disease), and current medication use. The tool had questions for assessment of PAD including the use of ECQ and ABI. Participants were also evaluated for diabetes-related complications, including peripheral neuropathy, joint deformities, diabetic foot ulcers, and lower limb amputations.

### Data Management and Analysis

Data were collected electronically using KoBoToolbox (Cambridge, MA, USA). Upon completion of data collection, the dataset was exported to Microsoft Excel (Microsoft Corp., Redmond, WA, USA) for initial data cleaning and consistency checks. The cleaned dataset was then imported into Stata version 18 (StataCorp, College Station, TX, USA) for statistical analysis.

At the univariate level, continuous variables were assessed for normality and summarized as means with standard deviations (SD) where appropriate, while categorical variables were presented as frequencies and percentages. The prevalence of PAD was calculated as the proportion of participants with confirmed PAD among the total study population, with corresponding 95% confidence intervals (CIs) reported to indicate precision. Prevalence estimates were further stratified by key demographic and clinical characteristics.

Bivariate analysis was performed using Poisson regression with robust error variance to assess the association between each independent variable and PAD. Variables with a p-value ≤ 0.20 at the bivariate level were considered for inclusion in the multivariable model. Multivariable analysis was conducted using modified Poisson regression to estimate adjusted prevalence ratios (aPRs) and their corresponding 95% CIs. Variables with a p-value ≤ 0.05 in the final model were considered independently associated with PAD.

## Results

### Description of the participants

Of the 304 eligible participants approached, 300 consented to participate, yielding a response rate of 98.7%. The majority of participants were aged ≥50 years (65.3%), and females constituted 67.3% of the study population. Most respondents (68.3%) rarely or occasionally consumed fruits, while 55.7% reported sitting for 1–4 hours per day. Physical inactivity was common, with 53.8% of participants engaging in less than 150 minutes of moderate-intensity physical activity per week (Table 1).

**Table 1.**
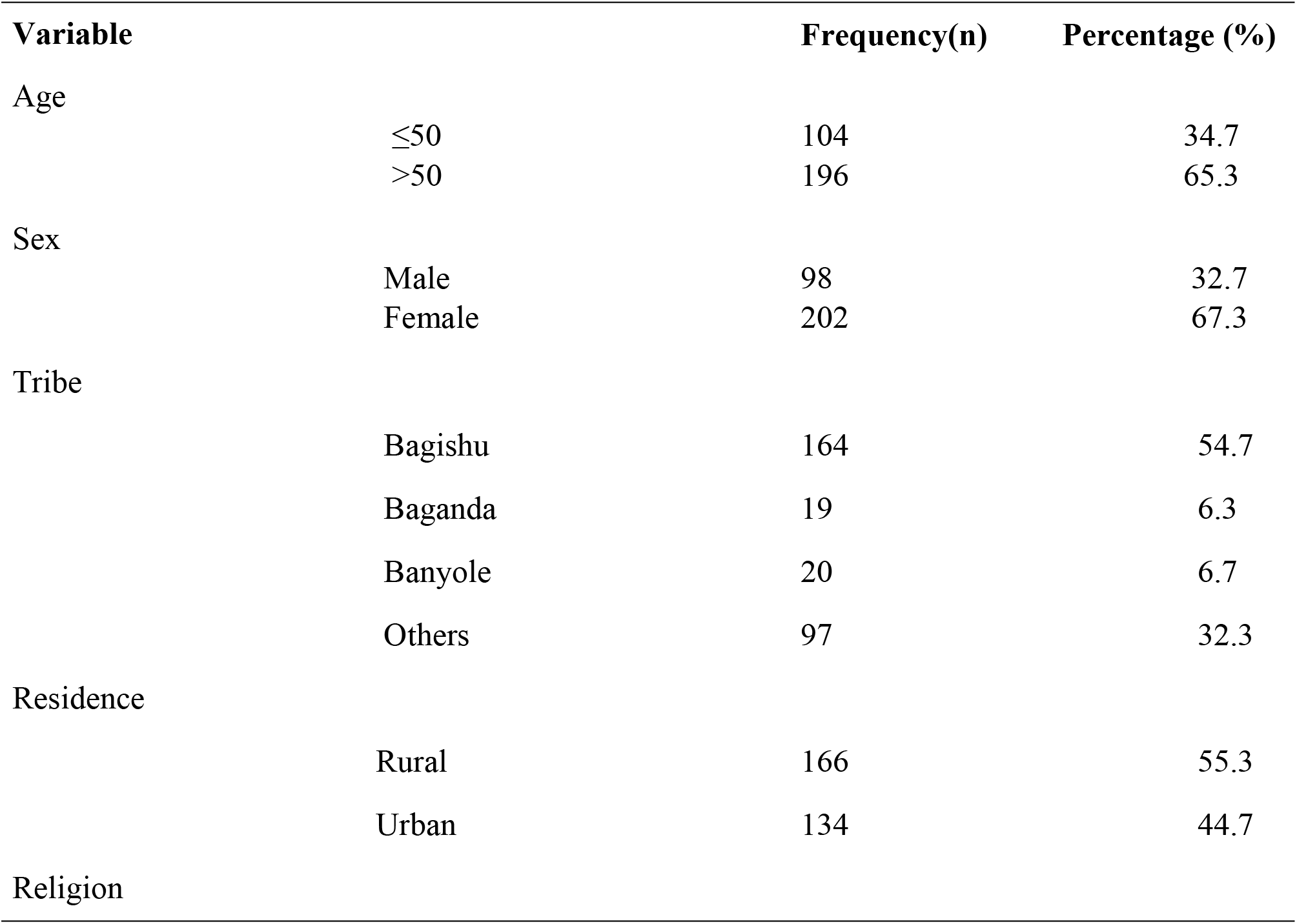
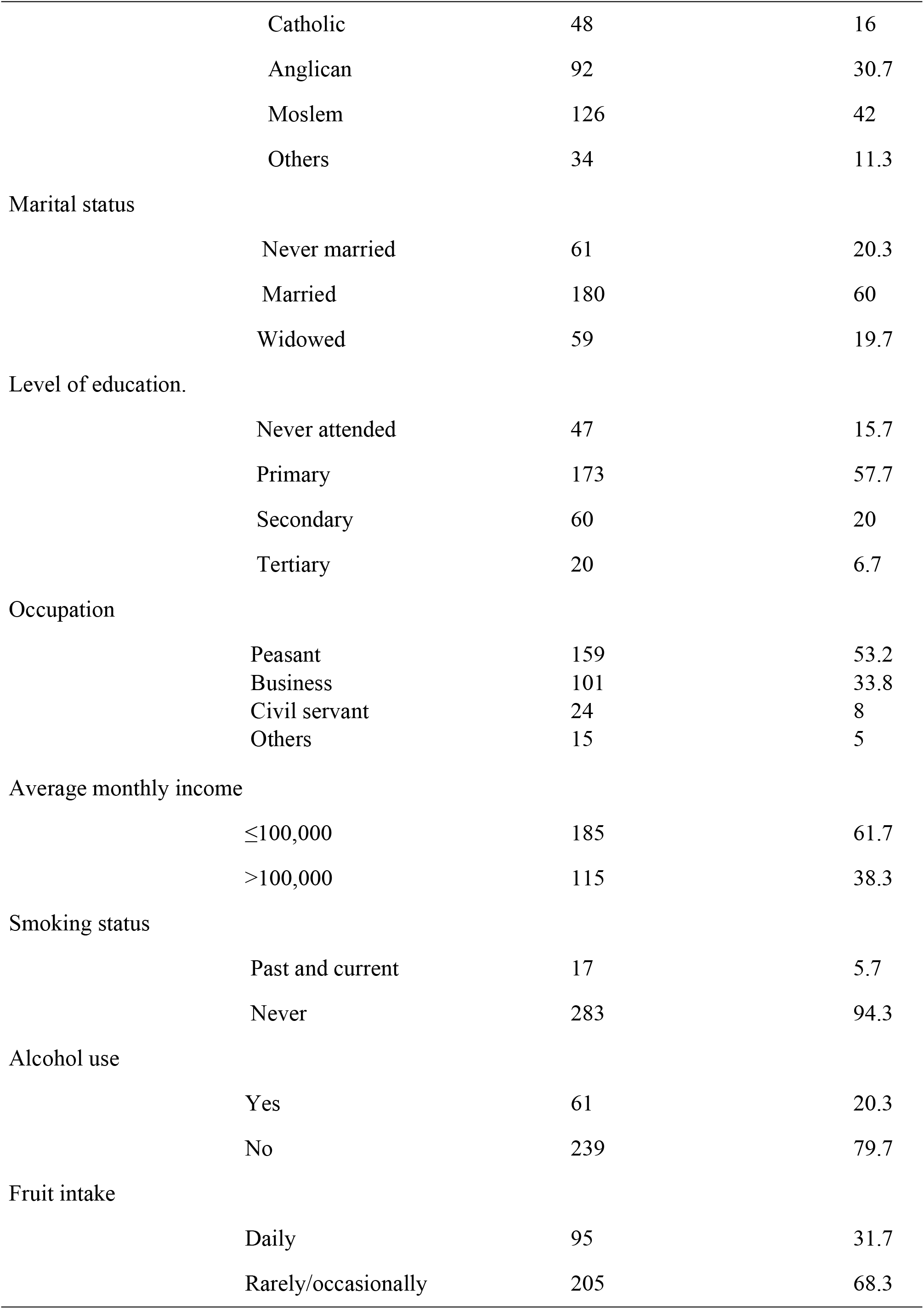
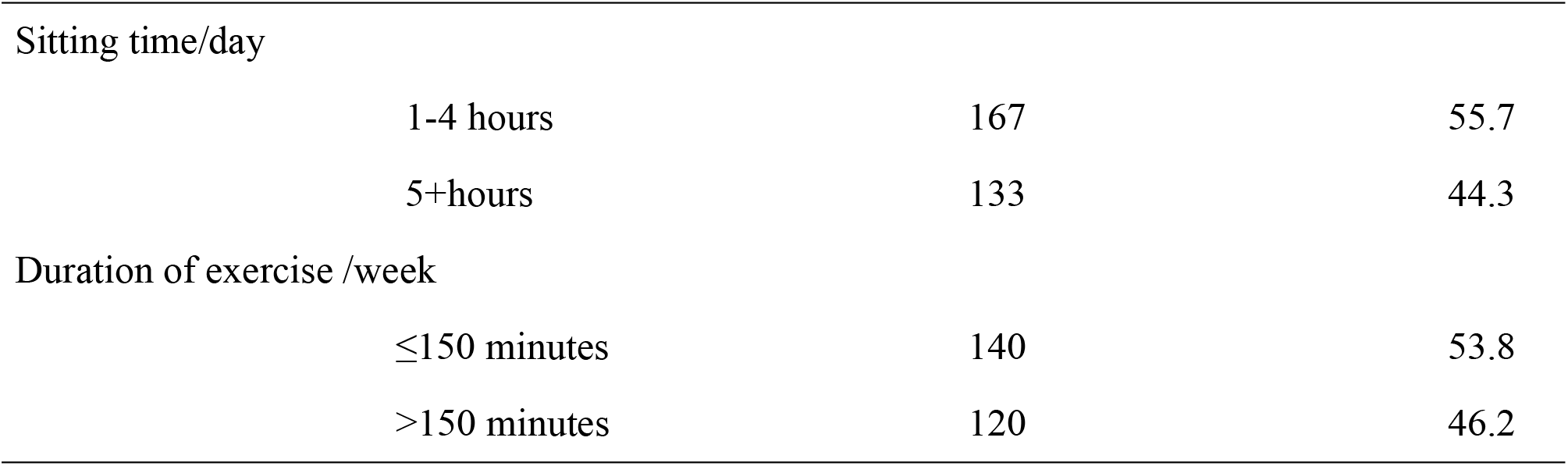
Characteristics of study participants.

| Variable | Frequency(n) | Percentage (%) |
| --- | --- | --- |
| Age |  |  |
| $\leq 50$ | 104 | 34.7 |
| $> 50$ | 196 | 65.3 |
| Sex |  |  |
| Male | 98 | 32.7 |
| Female | 202 | 67.3 |
| Tribe |  |  |
| Bagishu | 164 | 54.7 |
| Baganda | 19 | 6.3 |
| Banyole | 20 | 6.7 |
| Others | 97 | 32.3 |
| Residence |  |  |
| Rural | 166 | 55.3 |
| Urban | 134 | 44.7 |
| Religion |  |  |

|  |  |  |  |
| --- | --- | --- | --- |
|  | Catholic | 48 | 16 |
|  | Anglican | 92 | 30.7 |
|  | Moslem | 126 | 42 |
|  | Others | 34 | 11.3 |
| Marital status |  |  |  |
|  | Never married | 61 | 20.3 |
|  | Married | 180 | 60 |
|  | Widowed | 59 | 19.7 |
| Level of education. |  |  |  |
|  | Never attended | 47 | 15.7 |
|  | Primary | 173 | 57.7 |
|  | Secondary | 60 | 20 |
|  | Tertiary | 20 | 6.7 |
| Occupation |  |  |  |
|  | Peasant | 159 | 53.2 |
|  | Business | 101 | 33.8 |
|  | Civil servant | 24 | 8 |
|  | Others | 15 | 5 |
| Average monthly income |  |  |  |
|  | ≤100,000 | 185 | 61.7 |
|  | >100,000 | 115 | 38.3 |
| Smoking status |  |  |  |
|  | Past and current | 17 | 5.7 |
|  | Never | 283 | 94.3 |
| Alcohol use |  |  |  |
|  | Yes | 61 | 20.3 |
|  | No | 239 | 79.7 |
| Fruit intake |  |  |  |
|  | Daily | 95 | 31.7 |
|  | Rarely/occasionally | 205 | 68.3 |
| Sitting time/day |  |  |  |
|  | 1-4 hours | 167 | 55.7 |
|  | 5+hours | 133 | 44.3 |
| Duration of exercise /week |  |  |  |
|  | ≤150 minutes | 140 | 53.8 |
|  | >150 minutes | 120 | 46.2 |

### Participants’ comorbidities and medications

Overall, 38.3% of participants were overweight and 21.3% were classified as obese. Hypertension was present in 64.3% of the study population, with 51.3% exhibiting elevated blood pressure at the time of assessment. Poor glycemic control (HbA1c ≥ 7%) was observed in 77.7% of participants. Abnormal electrocardiographic findings were noted in 64.7% of the sample. Regarding lipid profiles, 59.7% of participants had elevated low-density lipoprotein (LDL) cholesterol, 44.3% had reduced high-density lipoprotein (HDL) cholesterol, and 40.0% had elevated total cholesterol levels. The use of antiplatelet agents and lipid-lowering medications was reported in 4.0% and 5.3% of participants, respectively (Table 2).

**Table 2.** Participant’s co-morbid conditions.

| Variable |  | Frequency(n) | Percentage (%) |
| --- | --- | --- | --- |
| Body mass index |  |  |  |
|  | Underweight | 24 | 8 |
|  | Normal | 97 | 32.3 |
|  | Overweight | 115 | 38.3 |
|  | Obese | 64 | 21.3 |
| DM of duration |  |  |  |
|  | <10 years | 229 | 76.3 |
|  | >10 | 71 | 23.7 |
| History of Hypertension |  |  |  |
|  | No | 107 | 35.7 |
|  | Yes | 193 | 64.3 |
| Blood pressure control |  |  |  |
|  | Normal | 146 | 48.7 |
|  | Elevated | 154 | 51.3 |
| Chronic kidney disease |  |  |  |
|  | No | 295 | 98.3 |
|  | Yes | 5 | 1.7 |
| Heart failure |  |  |  |
|  | No | 295 | 98.3 |
|  | Yes | 5 | 1.7 |
| HIV |  |  |  |
|  | No | 286 | 95.3 |
|  | Yes | 14 | 4.7 |
| Antiplatelet |  |  |  |
|  | No | 288 | 96 |
|  | Yes | 12 | 4 |
| Lipid-lowering drugs |  |  |  |
|  | No | 284 | 94.7 |
|  | Yes | 16 | 5.3 |
| Cardiac function |  |  |  |
|  | Normal ECG | 106 | 35.3 |
|  | Abnormal ECG | 194 | 64.7 |
| Glycemic control |  |  |  |
|  | Good (HbA1c <7) | 67 | 22.3 |
|  | Poor (HbA1c ≥7) | 233 | 77.7 |
| Total Cholesterol |  |  |  |
|  | Normal | 180 | 60 |
|  | High | 120 | 40 |
| HDL |  |  |  |
|  | Low | 133 | 44.3 |
|  | Normal | 167 | 55.7 |
| LDH |  |  |  |
|  | Optimal | 121 | 40.3 |
|  | High | 179 | 59.7 |
| Triglycerides to HDL ratio |  |  |  |
|  | <4 | 263 | 87.7 |
|  | ≥4 | 37 | 12.3 |

### Prevalence of Peripheral Artery Disease

Of the 300 participants, 42.3% (95% CI: 0.38–0.48) were diagnosed with PAD based on an abnormal ABI (ABI ≤ 0.9). Of these 15.8% had severe PAD disease (ABI <0.4). Using the ECQ, the prevalence of PAD was 37.0%, with 7.3% of participants reporting definite claudication (Table 3).

**Table 3.** Prevalence of PAD.

| PAD based on ABI | Frequency (N=300) | 95%CI |  | Percentage |
| --- | --- | --- | --- | --- |
| PAD | 127 | 0.38 | 0.48 | 42.3 |
| PAD based on ECQ |  |  |  |  |
| No claudication | 188 | 0.57 | 0.68 | 62.7 |
| Possible claudication | 90 | 0.25 | 0.35 | 30 |
| Definite claudication | 22 | 0.05 | 0.11 | 7.3 |
| Clinical severity of PAD (ABI) |  |  |  |  |
| Asymptomatic | 55 | 0.23 | 0.45 | 43.3 |
| Mild/moderate | 52 | 0.31 | 0.54 | 40.9 |
| Severe/rest pain | 20 | 0.16 | 0.36 | 15.8 |

**Table 4.** Bivariate analysis for Sociodemographic factors associated with PAD.

| Variable | No PAD<br>173 (57.7) | PAD present<br>127 (42.3) | CPR (95%CI) | P-Value |
| --- | --- | --- | --- | --- |
| **Age (Mean ± SD)** |  |  |  |  |
| | 53.66 ± 13.28 | 57.42±13.30 | 1.02 (1.00–1.04) | 0.071 |
| **Sex** |  |  |  |  |
| Male | 64 (37.0) | 34 (26.8) | 1 | 1 |
| Female | 109 (63.0) | 93 (73.2) | 1.48 (0.858 - 2.566) | 0.158 |
| **Tribe** |  |  |  |  |
| Bagishu | 103 (59.5) | 61 (48.0) | 1 | 1 |
| Baganda | 11 (6.4) | 8 (6.3) | 0.78 (0.241 - 2.559) | 0.688 |
| Banyole | 10 (5.8) | 10 (7.9) | 1.49 (0.625 - 3.558) | 0.368 |
| Others | 49 (28.3) | 48 (37.8) | 1.38 (0.832 - 2.3) | 0.211 |
| **Residence** |  |  |  |  |
| Rural | 94 (54.3) | 72 (56.7) | 1 | 1 |
| Urban | 79 (45.7) | 55 (43.3) | 1.2 (0.751 - 1.929) | 0.442 |
| **Religion** |  |  |  |  |
| Catholic | 29 (16.8) | 19 (15.0) | 1 | 1 |
| Anglican | 51 (29.5) | 41 (32.3) | 1.04 (0.488 - 2.229) | 0.912 |
| Moslem | 74 (42.8) | 52 (40.9) | 1.3 (0.64 - 2.622) | 0.472 |
| Others | 19 (11.0) | 15 (11.8) | 0.71 (0.241 - 2.065) | 0.525 |
| **Level of education.** |  |  |  |  |
| Never attended | 24 (13.9) | 23 (18.1) | 1 | 1 |
| Primary | 105 (60.7) | 68 (53.5) | 0.79 (0.423 - 1.491) | 0.473 |
| Secondary | 32 (18.5) | 28 (22.0) | 0.9 (0.43 - 1.9) | 0.79 |
| Tertiary | 12 (6.9) | 8 (6.3) | 0.54 (0.155 - 1.903) | 0.339 |
| **Occupation** |  |  |  |  |
| Peasant | 90 (52.3) | 69 (54.3) | 1 | 1 |
| Business | 63 (36.6) | 38 (29.9) | 0.91 (0.539 - 1.541) | 0.729 |
| Civil servant | 11 (6.4) | 13 (10.2) | 0.7 (0.249 - 1.954) | 0.493 |
| Others | 8 (4.7) | 7 (5.5) | 1.39 (0.549 - 3.543) | 0.484 |
| **Average monthly income** |  |  |  |  |
| ≤100,000 | 102 (59.0) | 83 (65.4) | 1 | 1 |
| >100,000 | 71 (41.0) | 44 (34.6) | 0.66 (0.39 - 1.105) | 0.113 |
| **Smoking status** |  |  |  |  |
| Past and current | 10 (5.8) | 7 (5.5) | 1 | 1 |
| Never | 163 (94.2) | 120 (94.5) | 1.32 (0.416 - 4.203) | 0.637 |
| **Alcohol use** |  |  |  |  |
| Yes | 38 (22.0) | 23 (18.1) | 1 | 1 |
| No | 135 (78.0) | 104 (81.9) | 2.26 (1.035 - 4.939) | 0.041 |
| **Fruit intake** |  |  |  |  |
| Daily | 48 (27.7) | 47 (37.0) | 1 | 1 |
| Rarely/occasionally | 125 (72.3) | 80 (63.0) | 0.99 (0.597 - 1.643) | 0.969 |
| **Sitting time/day** |  |  |  |  |
| 1-4 hours | 102 (59.0) | 65 (51.2) | 1 | 1 |
| 5+hours | 71 (41.0) | 62 (48.8) | 1.37 (0.854 - 2.197) | 0.192 |
| **Exercise /week** |  |  |  |  |
| ≤ 150 minutes | 75 (50.7) | 65 (58.0) | 1 | 1 |
| >150 minutes | 73 (49.3) | 47 (42.0) | 0.76 (0.45 - 1.288) | 0.309 |

**Table 5.** Bivariate analysis for comorbidities and medications associated with PAD.

| **Variable** | **No PAD**<br>**173 (57.7)** | **PAD**<br>**Present**<br>**127 (42.3)** | **CPR (95%CI)** | **p-**<br>**Value** |
| --- | --- | --- | --- | --- |
| Body mass index |  |  |  |  |
| Underweight | 13 (7.5) | 11 (8.7) | 1 | 1 |
| Normal | 56 (32.4) | 41 (32.3) | 0.91 (0.539 - 1.541) | 0.729 |
| Overweight | 67 (38.7) | 48 (37.8) | 0.7 (0.246 - 1.954) | 0.493 |
| Obese | 37 (21.4) | 27 (21.3) | 1.39 (0.548 - 3.542) | 0.484 |
| DM duration (Mean ± SD) | 6.24 ± 6.07 | 6.87 ± 5.38 | 1.01 (0.98–1.05) | 0.499 |
| Hypertension |  |  |  |  |
| No | 65 (37.6) | 42 (33.1) | 1 | 1 |
| Yes | 108 (62.4) | 85 (66.9) | 1.27 (0.759 - 2.116) | 0.365 |
| Blood pressure control |  |  |  |  |
| Normal | 88 (50.9) | 58 (45.7) | 1 | 1 |
| Elevated | 85 (49.1) | 69 (54.3) | 1.73 (0.927 - 3.219) | 0.085 |
| Chronic kidney disease |  |  |  |  |
| No | 170 (98.3) | 125 (98.4) | 1 | 1 |
| Yes | 3 (1.7) | 2 (1.6) | 0.87 (0.12 - 6.249) | 0.888 |
| HIV |  |  |  |  |
| No | 166 (96.0) | 120 (94.5) | 1 | 1 |
| Yes | 7 (4.0) | 7 (5.5) | 0.93 (0.292 - 2.953) | 0.9 |
| Antiplatelet |  |  |  |  |
| No | 169 (97.7) | 119 (93.7) | 1 | 1 |
| Yes | 4 (2.3) | 8 (6.3) | 1.88 (0.755 - 4.659) | 0.176 |
| lipid-lowering drugs |  |  |  |  |
| No | 167 (96.5) | 117 (92.1) | 1 | 1 |
| Yes | 6 (3.5) | 10 (7.9) | 1.69 (0.732 - 3.906) | 0.219 |
| Cardiac function |  |  |  |  |
| Normal ECG | 63 (36.4) | 43 (33.9) | 1 | 1 |
| Abnormal ECG | 110 (63.6) | 84 (66.1) | 0.85 (0.524 - 1.378) | 0.51 |
| Glycemic control (Mean ± SD) | | | | |
| | 10.18 ± 3.66 | 10.26 ± 3.18 | 1.00 (0.94–1.07) | 0.884 |
| Total Cholesterol (Mean ± SD) | | | | |
| | 4.85 ± 1.28 | 5.22 ± 1.74 | 1.14 (0.98–1.31) | 0.085 |
| HDL |  |  |  |  |
| Low | 76 (43.9) | 57 (44.9) | 1 | 1 |
| Normal | 97 (56.1) | 70 (55.1) | 0.82 (0.511 - 1.314) | 0.409 |
| LDH |  |  |  |  |
| Optimal | 73 (42.2) | 48 (37.8) | 1 | 1 |
| High | 100 (57.8) | 79 (62.2) | 1.44 (0.87 - 2.396) | 0.155 |
| Triglycerides to HDL ratio |  |  |  |  |
| <4 | 153 (88.4) | 110 (86.6) | 1 | 1 |
| ≥4 | 20 (11.6) | 17 (13.4) | 1.07 (0.529 - 2.148) | 0.858 |

### Factors associated with PAD

Participants with PAD were, on average, older than those without PAD (57.4 vs. 53.7 years). Each additional year of age was associated with an approximately 2% higher likelihood of PAD; however, this did not reach statistical significance (cPR 1.02, 95% CI: 1.00–1.04; p = 0.071).

The mean duration of diabetes was slightly longer among participants with PAD compared to those without PAD (6.9 vs. 6.2 years), although this difference was not statistically significant (cPR 1.01, 95% CI: 0.98–1.05; p = 0.49). Similarly, those with PAD had higher mean total cholesterol levels (5.22 ± 1.74 mmol/L) compared to those without PAD (4.85 ± 1.28 mmol/L), corresponding to a 14% higher likelihood of PAD per unit increase in cholesterol; this association did not reach statistical significance (cPR 1.14, 95% CI: 0.98–1.31; p = 0.09). Mean HbA1c levels were high in both groups (10.18% vs. 10.26%), indicating generally poor glycemic control in this population.

### 4.8 Multivariable Analysis of Factors Associated with Peripheral Artery Disease

Table 6 presents the multivariate results for factors associated with PAD. Participants older than 50 years had 1.24 times higher likelihood of developing PAD compared to those younger than 50 years (aPR 1.24, 95% CI: 0.73 - 2.09), though this association did not reach statistical significance (p=0.43). Female sex was associated with increased likelihood of PAD (aPR 1.46, 95% CI: 0.91–2.56), but the relationship was not statistically significant (p=0.18).

**Table 6.** Multivariable analysis.

| Variable | APR (95%CI) | p-value |
| --- | --- | --- |
| Age |  |  |
| >50 | 1.24 (0.73 - 2.09) | 0.43 |
| ≤50 | 1 | 1 |
| Sex |  |  |
| Female | 1.46 (0.84 - 2.55) | 0.18 |
| Male | 1 | 1 |
| Antiplatelet |  |  |
| Yes | 1.69 (0.66 - 4.29) | 0.27 |
|  | 1 | 1 |
| Cholesterol levels |  |  |
| Elevated | 1.39 (0.86 - 2.25) | 0.18 |
| Normal | 1 | 1 |
| Fruits intake |  |  |
| Rarely/occasionally | 1 | 1 |
| Daily | 1.02 (0.61 - 1.69) | 0.95 |
| Sitting time |  |  |
| ≥ hours | 1.28 (0.79 - 2.08) | 0.31 |
| < 5 hours | 1 | 1 |
| Glycemic control |  |  |
| Poor (HbA1c ≥7) | 1.35 (0.72 - 2.53) | 0.35 |
| Good (HbA1c <7) | 1 | 1 |

High total cholesterol was associated with 1.39 times higher chances of PAD relative to normal cholesterol levels; however, this did not achieve statistical significance (aPR 1.39, 95% CI: 0.86 - 2.25; p= 0.18).

There was no significant difference in the likelihood of PAD between participants who consumed fruits regularly and those who consumed fruits occasionally (aPR 1.02, 95% CI: 0.61 - 1.69, p=0.95). Sitting for five or more hours daily was associated with increased chances of PAD compared to sitting 1–4 hours (aPR 1.28, 95% CI: 0.79 - 2.08), without statistical significance (p=0.31). Poor glycemic control (HbA1c ≥7) was associated with a nonsignificant increase in the likelihood of PAD compared to good control (aPR 1.35, 95% CI: 0.72 - 2.53; p=0.35).

## Discussion

In this study, we found a high prevalence of PAD of 42.3% (95% CI: 38.0–48.0) based on ABI measurements and 37% (95% CI: 31.9 - 42.8) based on ECQ. The magnitude of PAD in this setting indicates a substantial burden of both asymptomatic and symptomatic atherosclerotic disease among adults with DM. This prevalence is notably higher than the 24% to 39% reported in southwestern Uganda and Central Uganda respectively [10, 18]. The observed difference may reflect variations in study populations, temporal changes in cardiovascular risk profiles, or differences in access to preventive and screening services. Notably, the prevalence observed in our study exceeds pooled estimates from meta-analyses conducted in low- and middle-income countries, which range from 20% to 35.7% [19, 20]. The high burden of PAD in this resource-limited setting underscores the potential for a disproportionately higher burden of atherosclerotic disease among patients with DM in resource-limited settings. This underscores the need to routinize intensified screening and prevention efforts for PAD among DM patients in low income countries. Addressing the high burden of PAD among DM patients requires changes to the health system with a focus on improving health literacy, better healthcare-seeking behaviour, improved routine screening for PAD, and implementation of targeted primary prevention interventions.

Unlike previous studies, we did not find a significant association between PAD and advancing age, hypertension, poor glycemic control, dyslipidemia, and physical inactivity [21-23]. This finding is probably due to lack of statistical power, given the smaller sample size. Furthermore, the influence of unmeasured confounders such as medication adherence, access to care, and genetic susceptibility cannot be excluded. Measurement error, particularly in self-reported behavioral variables, may also have attenuated observed associations. Nevertheless, a large proportion of participants had a low fruit consumption (68.3%), insufficient physical activity (53.8% reporting <150 minutes of exercise per week), elevated total cholesterol (40%), and high low-density lipoprotein (LDL) levels (59.7%). This indicates poor behavioral and lifestyle practices among patients with DM, which may likely predispose them to long-term complications including PAD.

Overall, while conventional cardiovascular risk factors demonstrated trends consistent with prior evidence, their lack of statistically significant associations in this study highlights the potential role of contextual or unmeasured determinants of PAD in this population. These findings underscore the need for larger, longitudinal studies incorporating comprehensive risk profiling to better elucidate the drivers of PAD among patients with DM in similar settings.

## Strengths and Limitations

This study provides important insights into the burden and correlates of PAD among adults with DM in a resource-limited setting. A key strength is the use of the ABI, a validated and objective diagnostic tool that enables detection of both symptomatic and asymptomatic PAD, thereby minimizing misclassification bias [24]. In addition, standardized measurement procedures and the use of trained personnel enhanced the reliability and reproducibility of the data collected. The study also achieved a high response rate which reduced the likelihood of selection bias.

However, several limitations should be acknowledged. The cross-sectional design precludes causal inference; thus, observed relationships should be interpreted as associations rather than evidence of causality. Some exposure variables, including smoking status, dietary patterns, and physical activity, were based on self-report and are therefore subject to recall and social desirability bias. Residual cofounding cannot be excluded, as certain relevant variables such as medication adherence and genetic susceptibility were not measured. The statistical power in the study may have been low to detect modest associations within multivariable analyses.

## Conclusion

The prevalence of PAD was high in our setting where close to a half (42.3%) of patients with DM had PAD. Given the high prevalence of asymptomatic and undiagnosed PAD among adults with DM, emphasis should be placed on patient education regarding clinical symptoms and complications of PAD to promote a healthy lifestyle, early recognition and management of PAD. Clinicians should be encouraged to conduct routine screening using simple, low-cost tools such as vascular Doppler for ABI measurement, particularly in settings with limited access to specialized vascular care. Efforts should focus on integrating PAD screening protocols into national diabetes management guidelines and ensuring the availability of basic diagnostic tools at primary healthcare facilities. The high burden of PAD amidst poor lifestyle practices reinforces the rationale for comprehensive cardiovascular risk management in diabetes, addressing dyslipidemia, glycaemic control, physical inactivity, and sedentary behaviours. Strengthening integrated diabetes care bundle to detect and manage PAD early remains a prudent public health priority.

## Data Availability

The dataset generated and analyzed during the current study contains potentially identifiable participant information. In accordance with the conditions of the ethical approval granted by the Institutional Review Board and the informed consent provided by participants, the data cannot be deposited in a public repository. De-identified data may be made available to qualified researchers upon reasonable request to the corresponding author, subject to approval by the relevant ethics committee and institutional requirements to protect participant confidentiality.

## Acknowledgements

We thank the management and staff of Mbale Regional Referral Hospital, Busitema University and the participants for their respective contribution in the implementation of the study.

## Funding statement

Research was supported by the Department of Internal Medicine Busitema University, the Department of Physiology Busitema University, Uganda Diabetic Association, and Mbale Regional Referral Hospital.

## Conflict of interest

The authors declare that they have no conflict of interest.

## Declarations

### Ethical approval and consent to participate

Ethical clearance was obtained from Busitema University Faculty of Health Sciences Institutional Review Board (BUFHS IRB), approval number [BUFHS-2024-232]. All data and study documents were stored securely, according to Good Clinical Practice, NIDA’s Version 5 on secure storage of research materials [25]. Participants provided written informed consent before enrolling in the study.

### Consent for publication

Not applicable

### Contributor Information

JI, DB, DI, and AM were involved in the design, and conceptualization of the study. JI, ON, JK, EK, and EA participated in data collection including physical assessment of the participants. DB, AM and DI supervised the overall implementation of the study. JI and JE were involved in data analysis. JI wrote the first draft of the manuscript, while ON, JK, EA, PMM, EK, RK, AM, DI, DB reviewed the draft manuscript. All authors have met the criteria of authorship, and have read and consented for the paper to be submitted for publication.

## Notes

### Competing Interest Statement

The authors have declared no competing interest.

### Funding Statement

The author(s) received no specific funding for this work.

### Author Declarations

Ethical clearance was obtained from Busitema University Faculty of Health Sciences Institutional Review Board (BUFHS IRB), approval number [BUFHS-2024-232].

## References

1. Criqui MH, Matsushita K, Aboyans V, Hess CN, Hicks CW, Kwan TW, et al. Lower extremity peripheral artery disease: contemporary epidemiology, management gaps, and future directions: a scientific statement from the American Heart Association. Circulation. 2021;144(9):e171–e91.

2. Soyoye DO, Abiodun OO, Ikem RT, Kolawole BA, Akintomide AO. Diabetes and peripheral artery disease: A review. World journal of diabetes. 2021;12(6):827.

3. Huijberts MS, Schaper NC, Schalkwijk CG. Advanced glycation end products and diabetic foot disease. Diabetes/metabolism research and reviews. 2008;24(S1):S19–S24.

4. Edmonds M, Manu C, Vas P. The current burden of diabetic foot disease. Journal of clinical orthopaedics and trauma. 2021;17:88–93.

5. Kohn CG, Alberts MJ, Peacock WF, Bunz TJ, Coleman CI. Cost and inpatient burden of peripheral artery disease: Findings from the National Inpatient Sample. Atherosclerosis. 2019;286:142–6.

6. Shaw JE, Sicree RA, Zimmet PZ. Global estimates of the prevalence of diabetes for 2010 and 2030. Diabetes research and clinical practice. 2010;87(1):4–14.

7. GBD. Global, regional, and national burden of diseases and injuries for adults 70 years and older: systematic analysis for the Global Burden of Disease 2019 Study. bmj. 2022;376.

8. Shamaki GR, Markson F, Soji-Ayoade D, Agwuegbo CC, Bamgbose MO, Tamunoinemi B-M. Peripheral artery disease: a comprehensive updated review. Current Problems in Cardiology. 2022;47(11):101082.

9. Okello S, Millard A, Owori R, Asiimwe SB, Siedner MJ, Rwebembera J, et al. Prevalence of lower extremity peripheral artery disease among adult diabetes patients in southwestern Uganda. BMC Cardiovascular Disorders. 2014;14:1–6.

10. Mwebaze RM, Kibirige D. Peripheral arterial disease among adult diabetic patients attending a large outpatient diabetic clinic at a national referral hospital in Uganda: a descriptive cross sectional study. PLoS One. 2014;9(8):e105211.

11. Johnston LE, Stewart BT, Yangni-Angate H, Veller M, Upchurch GR, Gyedu A, et al. Peripheral arterial disease in sub-Saharan Africa: a review. JAMA surgery. 2016;151(6):564–72.

12. Akalu Y, Birhan A. Peripheral arterial disease and its associated factors among type 2 diabetes mellitus patients at Debre Tabor general hospital, Northwest Ethiopia. Journal of diabetes research. 2020;2020.

13. Okello S, Millard A, Owori R, Asiimwe SB, Siedner MJ, Rwebembera J, et al. Prevalence of lower extremity peripheral artery disease among adult diabetes patients in southwestern Uganda. BMC Cardiovascular Disorders. 2014;14(1):1–6.

14. Song P, Rudan D, Zhu Y, Fowkes FJ, Rahimi K, Fowkes FGR, et al. Global, regional, and national prevalence and risk factors for peripheral artery disease in 2015: an updated systematic review and analysis. The Lancet Global Health. 2019;7(8):e1020–e30.

15. Wang Z, Wang X, Hao G, Chen Z, Zhang L, Shao L, et al. A national study of the prevalence and risk factors associated with peripheral arterial disease from China: The China Hypertension Survey, 2012–2015. International journal of cardiology. 2019;275:165–70.

16. Okello S, Millard A, Owori R, Asiimwe SB, Siedner MJ, Rwebembera J, et al. Prevalence of lower extremity peripheral artery disease among adult diabetes patients in southwestern Uganda. BMC Cardiovasc Disord. 2014;14:75. Epub 2014/06/11. doi: 10.1186/1471-2261-14-75. PubMed PMID: 24913468; PubMed Central PMCID: PMCPMC4057935.

17. Firnhaber JM, Powell C. Lower extremity peripheral artery disease: diagnosis and treatment. American family physician. 2019;99(6):362–9.

18. Okello S, Millard A, Owori R, Asiimwe SB, Siedner MJ, Rwebembera J, et al. Prevalence of lower extremity peripheral artery disease among adult diabetes patients in southwestern Uganda. BMC Cardiovascular Disorders. 2014;14(1):75.

19. Song P, Zhang Y, Yu J, Zha M, Zhu Y, Rahimi K, et al. Global Prevalence of Hypertension in Children: A Systematic Review and Meta-analysis. JAMA Pediatr. 2019;173(12):1154–63. doi: 10.1001/jamapediatrics.2019.3310. PubMed PMID: 31589252; PubMed Central PMCID: PMCPMC6784751.

20. Haile KE, Amsalu A, Kassie GA, Asgedom YS, Azeze GA. Exploring the Prevalence and Risk Factors of Peripheral Artery Disease in Type 2 Diabetes Patients in Sub-Saharan Africa: A Systematic Review and Meta-analysis. Frontiers in Clinical Diabetes and Healthcare. 2025;6:1563984.

21. Narres M, Claessen H, Droste S, Kvitkina T, Koch M, Kuss O, et al. The Incidence of End-Stage Renal Disease in the Diabetic (Compared to the Non-Diabetic) Population: A Systematic Review. PLoS One. 2016;11(1):e0147329. Epub 20160126. doi: 10.1371/journal.pone.0147329. PubMed PMID: 26812415; PubMed Central PMCID: PMCPMC4727808.

22. Criqui MH, Aboyans V. Epidemiology of peripheral artery disease. Circ Res. 2015;116(9):1509–26. doi: 10.1161/circresaha.116.303849. PubMed PMID: 25908725.

23. Peer N, Kengne A-P, Motala AA, Mbanya JC. Diabetes in the Africa Region: an update. Diabetes research and clinical practice. 2014;103(2):197–205.

24. Gerhard-Herman MD, Gornik HL, Barrett C, Barshes NR, Corriere MA, Drachman DE, et al. 2016 AHA/ACC Guideline on the Management of Patients With Lower Extremity Peripheral Artery Disease: A Report of the American College of Cardiology/American Heart Association Task Force on Clinical Practice Guidelines. Circulation. 2017;135(12):e726–e79. Epub 20161113. doi: 10.1161/CIR.0000000000000471. PubMed PMID: 27840333; PubMed Central PMCID: PMCPMC5477786.

25. NIDA. Good Clinical Practices 2021 [cited 2024]. Available from: https://gcp.nidatraining.org/changelog.

